# Assessing medication-related burden and medication adherence among older patients from Central Nepal: A machine learning approach

**DOI:** 10.64898/2026.04.22.26351447

**Authors:** Roshan Giri, Rohit Agrawal, Sabin Raj Lamichhane, Sachita Barma, Rachana Mahatara

## Abstract

**Background:** Nepal is experiencing a rapid demographic shift toward an aging population, with concurrent increase in morbidity and medication-related problems. Despite this, the multidimensional experience of medication-related burden (MRB) and refill adherence remain under-studied, particularly through the lens of socio-demographic, clinical and medication-related predictive features. This study aimed to assess MRB and medication adherence, and utilize machine learning (ML) architectures to identify complex factors influencing both.

**Methods:** A cross-sectional study conducted among 390 ambulatory older patients (aged≥65 years) at Bharatpur Hospital, Nepal. MRB and medication adherence was assessed using Living with Medications Questionnaire (LMQ-3) and Adherence to Refills and Medication Scale (ARMS). Six ML architectures (Ordinary Least Square, LightGBM, Random Forest, XGBoost, SVM, and Penalized linear regression) were employed to predict ARMS and LMQ scores using various socio-demographic, clinical and medication-related predictive features. Model explainability was provided through SHAP (Shapley Additive exPlanations). All the analysis were performed using R.

**Results:** The median LMQ-3 score was 110.0 (IQR 14.0), reflecting a moderate medication-related burden, while the median ARMS score of 21.0 (IQR 6.0) indicated moderate non-adherence. Random forest was the superior predictive model for both MRB and adherence. SHAP analysis revealed requiring assistance for medication and polypharmacy as the most significant drivers of both increased burden and poor adherence. Interaction analysis revealed that while polypharmacy typically worsens adherence, the risk is partially mitigated when patients receive physical or cognitive assistance. Additional, financial factors and employment status emerged as significant predictors.

**Conclusion:** Older patients in Nepal face a significant medication-related burden and non-adherence, driven largely by regimen complexity and the need for support. The high predictive accuracy of ML models suggests that clinical interventions should prioritize simplified regimens and patient-centered counseling for those with high dependency. These findings provide a data-driven rational for policy-level medication optimization strategies in Nepal’s evolving healthcare system.

**Author Summary:** As ageing occurs, there is high chance of presence of the chronic conditions. For management of these conditions, older people are often prescribed with multiple medications and often are vulnerable to those medications whose risk outweighs benefits. As a result there is high chance of occurrence of adverse effects; these effects have caused substantial degradation of the health related quality of life. Although pharmacotherapy is the mainstay of the chronic disease management, older people often feel medication burden. Medication burden is practical experience arising from the practical and psychological challenge while managing medications. Research shows high burden causes the non-adherence, a significant problem among older adults, causing significant problem in pharmacotherapy. Hence, we used validated Questionnaire for assessing the medication burden and medication adherence among older ambulatory adults attending the central hospital of Nepal. We used machine learning approach for the high prediction of the predictors influencing the medication related burden and mediation adherence. Moderate burden was observed among older adults and moderate non-adherence was also observed. We found needing assistance for medication management and multiple medications were the strongest predictors for both Medication burden and non-adherence. Our Study provides new insights and area for the implementation of clinical intervention for the medication optimization.

## Introduction

Global ageing estimates suggests that by 2050, two-thirds of the population aged over 60 will reside in low- and middle income countries (1). This trend is evident in Nepal; the 2021 census revealed that 5.90% of population is aged 65 or older, a significant increase from 3.29% in 2011 (2). This demographic shift is accompanied by a high prevalence of non-communicable chronic conditions, with nearly 48.9% of older Nepalese individuals living with at least one such condition (3). Managing theses chronic conditions require long-term pharmacotherapy, which is inherently challenging due to multimorbidity and age-related changes in pharmacokinetic and Pharmacodynamic profiles (4). Older patients are frequently exposed to polypharmacy and potentially inappropriate medications (PIMs), increasing the risk of adverse drug reactions and drug interactions (5). These events subsequently lead to prolonged hospital stays, increased medication-related burden, decreased health-related quality of life (HRQoL) and higher healthcare costs (6–8). In Nepal, more than 33% of ambulatory older patients receive PIMs, and over 40% are non-adherent to their therapy (9,10).

Medication-related burden (MRB) is a negative experience arising from the practical and psychological challenges of managing medications (11,12), as well as the impact of healthcare interventions. Evidences shows that MRB plays a crucial role in affecting patients’ well-being, beliefs and behaviors (13). High MRB results in substantial consequences: non-adherence, poor clinical outcomes, decreased HRQoL, decreased trust in providers, and psychological dissatisfaction. Previous studies have shown numerous factors; including medication regimen, social factors, communication with health care provider, side effects and health care cost (14), to affect MRB. Systematic reviews have revealed that many patients hold negative views towards medication, while some refusing use due to concerns regarding adverse reactions and the risk of dependency (13).

Various strategies are employed for medication optimization, such as deprescribing and medication reviews (15,16); however, patient perception on MRB is often under-looked. Research in Nepal indicates that over half of the patients are willing to have their medication de-prescribed, suggesting a desire to reduce burden (5). Patient experience is significantly shaped by the quality of the therapeutic relationship; specifically, a positive rapport with clinicians and a clear understanding of benefits have shown to ease restrictive medication taking-beliefs. Therefore, investing the lived experience of patients on long-term medications is essential for achieving effective therapeutic management. While existing qualitative and quantitative researches have explored various facets of medications beliefs, satisfaction, and management capacity, with or without focus on particular medical condition or medication therapy, there remains a need for a comprehensive assessment of the overall burden (17). Furthermore, in the context of Nepal’s evolving healthcare system, where older patients face varying co-payment structures, understanding the interaction between economic factors and perceived burden is vital.

While several tools exists to measure MRB, such as Medication Regimen Complexity Index, or the Drug Burden Index, the Living with Medicines (LMQ-3) questionnaire. is uniquely patient-centered, comprehensive and multidimensional, covering all domains that could be burdensome to the individual (18–20). However, the factors influencing MRB and its subsequent effect on medication refills are often complex and non-linear. Traditional statistical methods may fail to capture the intricate patterns between socio-demographic, clinical, and medication-related variables (21). Machine learning (ML) offers a robust framework for identifying these non-linear interactions and providing superior predictive accuracy, which is essential for developed targeted clinical interventions (22). Furthermore, while the prevalence of non-adherence is documented, the specific interplay of socio-demographic, clinical and medication related factors that drive these behaviors in the Nepalese older population remain poorly understood.

Despite the known prevalence of chronic illness and medication-related problems in Nepal, there is a critical lack of evidence regarding how multidimensional socio-demographic, clinical and medication related factors influence medication-related burden and adherence behaviors among geriatric patients. Therefore, this study aimed to: (1) assess the medication-related burden from patient perspectives using LMQ-3; (2) assess adherence to medication refills using Adherence to Refills and Medications Scale (ARMS); and (3) to evaluate features influencing MRB and medication adherence using various machine learning algorithms.

## Materials and Methods

### Ethics statement

Ethical approval was taken from Bharatpur Hospital Institutional Review Board (Ref Num: 082/083-06). A written informed consent was obtained from each participant. Permission was taken from the developer of the tools used in this study. This study was reported using Transparent Reporting of a multivariable prediction model for Individual Prognosis or Diagnosis AI (TRIPOD+AI) checklist (S1 File) (23).

### Study Area

Nepal is a South Asian landlocked country with geographical area of 1, 47,181 sq. km and an estimated population of 31.2 million. In Nepal, health care system includes private and public sector (24). Government of Nepal has endorsed health insurance system which includes emergency care free of cost, while OPD service is 10% co-payable in public sector and 20% in private sector. Geriatrics population of aged 70 above is non-co-payable while those aged 65-69 are co payable. Primary care is delivered through the local levels distributed all over the 7 province. Secondary and tertiary healthcare is delivered through the tertiary health care providers.

### Participants’ enrollment

A descriptive cross sectional study was conducted at Bharatpur hospital, a Central Nepal tertiary public Hospital, which provides specialized geriatric care from its dedicated OPD unit. The hospital provides services to its nearby 10 districts and is one of the largest Central Hospital in central Nepal under ministry of Health and Population. The target population was geriatric patients aged ≥ 65 years using at least one prescribed medicine for any chronic condition for at least one year. Patient with psychiatric disorder or cognitive impairment were excluded from the study. Sample size was calculated a priori using the “pmsampsize” R package (version 1.1.3) for multivariable prediction modeling with continuous outcomes (25,26). With 17 candidate predictor features and a conservative anticipated Cox-Snell R² of 0.30, the calculation required a minimum of 372 participants with complete data to limit shrinkage of the apparent R² to ≤10% and ensure precise estimation of model performance and intercept (approximately 22 observations per predictor parameter). Allowing for an expected 5% rate of incomplete data in this geriatric outpatient cohort, the final sample size was set at 390 participants.

### Measurement tools and scoring

This study had two major outcome variables; medication-related burden assessed by LMQ-3 questionnaire, and medication adherence by ARMS scale. LMQ-3 questionnaire contains 41 items each with 5-points Likert scale (1=strongly agree to 5= strongly disagree for forward coded questionnaire and 1= strongly disagree to 5= strongly agree for reverse coded questionnaire) LMQ questionnaire. The 41 items are divided into eight domains: relationship with health care professional about medicines (5 items), practical difficulties (7 items), cost related burden (3 items), side effects burden of prescribed medication (4 items), perceived effectiveness of medicines (6 items), attitudes/concerns about medication use (7 items), interference with day to day life (6 items), and control/autonomy about medication use (three items). The questionnaire also contains self-reported Visual analogue scale (VAS) ranging from 0 to 10, for self-reporting the medication-related burden with higher score indicating higher burden. Domains score (Overall LMQ score) are then summed to calculate total MRB, which ranges from 41 to 205, with higher score indicating higher burden (13). ARMS scale contains 12 questionnaires to assess the refills adherence. Each item was rated using 4 point Likert scale, ranging from 1 (none of the times) to 4 (all of the times). The overall ARMS score ranged from 12 to 48 with low score indicating better adherence (27).

Both LMQ-3 questionnaire and ARMS scale were translated to Nepali following approval from developer of each tool. Two native Nepali-speaking pharmacists independently forward-translated the English instrument into Nepali (FT1, FT2); discrepancies were resolved by consensus to produce a reconciled version (FTR). This was back-translated into English by two independent blinded translators (one pharmacist, one professional translator) (BT1, BT2), then reconciled into a single back-translation (BTR). Cognitive debriefing with 25 older adults using think-aloud and probing techniques assessed clarity, comprehension, and cultural relevance; minor wording adjustments were made based on feedback, yielding the final Nepali LMQ-3 and ARMS questionnaire. The prepared draft was then piloted in 10% of the sample (not included in main study) to ensure its reliability. Since the English versions of the LMQ-3 and ARMS have previously demonstrated robust psychometric properties in diverse populations, and the primary objective of this study was exploratory assessment in a Nepali-speaking geriatric cohort, only forward-backward translation with cognitive debriefing was performed. Full psychometric validation was beyond the scope of this cross-sectional study.

### Data Collection and preprocessing

Data were collected via face to face interviews of the study participants in drug counseling unit of the Hospital Pharmacy. Patients visiting to hospital pharmacy and refill counter for their medication refilled were approached for the study. Upon receiving verbal and written consent, data were collected. Structure questionnaire was consisting sections for socio-demographic characteristics, clinical characteristics and medication related characteristics, along with translated LMQ-3 and ARMS questionnaire. Information on patient socio demographic characteristics was obtained through interviews, while clinical characters and details information on drug formulation, frequency were collected by reviewing their prescription.

Data pre-processing were done using MS Excel. A total of 17 features were identified as potential predictive features for the outcome (S2 Table). This included socio-demographic features, clinical features and medication related features. All the features were categorical in nature. Socio-demographics features were categorized using pre-specified levels based on established standards. Domain specific thresholds were applied for CCI based on validated cut-offs (28). Medication use duration was categorized using threshold derived from median. Categorical features with sparse distributions (less than 5% frequency) were collapsed to guarantee statistical robustness. Since there were no missing data in the study, all participants’ outcome were calculated completely. Using the **recipes** package (version 1.3.1) (29), data preprocessing was managed, where categorical features were transformed using dummy encoding.

### Data analysis

Descriptive analysis was performed for LMQ-3 score, LMQ VAS score, LMQ domain scores, ARMS score and predictor features. Categorical features were expressed in terms of percentage and frequencies. Internal consistency was determined by Cronbach’s α. Differences between LMQ-3 score, LMQ VAS score, LMQ-3 domain scores and ARMS score among participants’ features were determined using parametric and non-parametric test as needed.

Predictive modelling of LMQ-3 score and ARMS score was performed using a supervised machine learning framework in R (**tidymodels**) (29). To capture diversion clinical data patterns, five algorithmic architectures were evaluated: Penalized Linear Regression (LR), Random Forest (RF), Extreme Gradient Boosting (XGBoost), Light Gradient Boosting Machine (LightGBM), and Support Vector Machines (SVM). An Ordinary Least Squares (OLS) regression served as a traditional statistical baseline. The dataset was partitioned into an 80% training and a 20% hold-out test set using stratified sampling. A 10×10 repeated cross-validation and an ANOVA-based racing algorithm was employed for efficient hyperparameter optimization. Model performance was rigorously assessed via Root Mean Square Error (RMSE) and Mean Absolute Error (MAE), R-Squared (R^2^), Mean Absolute Percentage Error (MAPE) and Huber loss, with 95% confidence intervals calculated via bootstrapping. Model integrity was verified through a diagnostic suite assessing homoscedasticity (Breusch-Pagan test), functional form (Ramsey RESET), and residual independence (Durbin-Watson). Model “explainability” was performed using SHAP (SHapley Additive exPlanations) values to rank feature importance and visualize clinical “tipping points” via dependence plots. Finally, bootstrapped OLS regression coefficients (or Odds Ratios for classification) were calculated to provide stable clinical effect sizes. A detailed description of the modeling pipeline, software versions, and diagnostic plots; along with results of hyperparameter optimization are available in S2 File. The results of diagnostic tests are present in S3 File and S4 File.

## Results

### Participant demographics

A total of 390 older adults (aged ≥65 years) visiting the outpatient pharmacy were enrolled. Two-thirds (64.9%) were young-old (65–74 years), with over half being female (55.4%). The majority resided inside Chitwan district (69.0%), was married (67.4%), unemployed (59.0%), illiterate (68.2%), of Brahmin ethnicity (58.2%), and reported medium economic status (85.9%). Common primary diagnoses included type 2 diabetes mellitus (31.3%), hypertension (29.2%), and chronic obstructive pulmonary disease (18.5%). Multimorbidity was prevalent (77.4%), with moderate Charlson Comorbidity Index category most frequent (64.4%). Polypharmacy affected 70.8%, predominantly with oral-only formulations (68.5%) and twice-daily frequency (63.8%). Assistance with medications was required by 30.5%, out-of-pocket payments by 41.3%, and short-duration medication use (≤10 years) by 56.7% (Table 1).

**Table 1.** Demographic and medication-related characteristics of participants (N = 390)

| **Characteristics** |  | **N (%)** |
| --- | --- | --- |
| *Age Group* | Young-old | 253 (64.9%) |
|  | Old-old | 135 (35.1%) |
| *Gender* | Female | 216 (55.4%) |
|  | Male | 174 (44.6%) |
| *Residence* | Inside Chitwan | 269 (69.0%) |
|  | Out of Chitwan | 121 (31.0%) |
| *Marital Status* | Married | 263 (67.4%) |
|  | Non-married | 127 (32.6%) |
| *Occupation* | Unemployed | 230 (59.0%) |
|  | Retired | 85 (21.8%) |
|  | Employed | 75 (19.2%) |
| *Educational Status* | Illiterate | 266 (68.2%) |
|  | Primary | 100 (25.6%) |
|  | Post-primary | 24 (6.2%) |
| *Ethnicity* | Brahamin | 227 (58.2%) |
|  | Janajati | 77 (19.7%) |
|  | Chhetri | 55 (14.1%) |
|  | Others | 31 (7.9%) |
| *Economic Status* | Medium | 335 (85.9%) |
|  | Low | 55 (14.1%) |
| *Primary Diagnosis* | T2DM | 122 (31.3%) |
|  | HTN | 114 (29.2%) |
|  | COPD | 72 (18.5%) |
|  | IHD | 40 (10.3%) |
|  | Others | 42 (10.8%) |
| *Multi-morbidity* | Yes | 302 (77.4%) |
|  | No | 88 (22.6%) |
| *CCI Category* | Mild | 24 (6.2%) |
|  | Moderate | 251 (64.4%) |
|  | Severe | 115 (29.5%) |
| *Polypharmacy* | Yes | 276 (70.8%) |
|  | No | 114 (29.2%) |
| *Medication Formulation* | Oral only | 267 (68.5%) |
|  | Oral + Non-oral | 123 (31.5%) |
| *Medication Frequency* | Twice Daily | 249 (63.8%) |
|  | Once Daily | 141 (36.2%) |
| *Requires Assistance* | No | 271 (69.5%) |
|  | Yes | 119 (30.5%) |
| *Pays for Medications* | No | 229 (58.7%) |
|  | Yes | 161 (41.3%) |
| *Medication Use Duration* | Short | 221 (56.7%) |
|  | Long | 169 (43.3%) |

### Medication-related burden (LMQ Score)

The median total Living with Medicines Questionnaire (LMQ-3) score was 110.0 (IQR 14.0; range 84–141), indicating moderate overall medication-related burden, with a median visual analog scale score of 4.0 (IQR 2.0; range 1–8) (Table 2). Item-level responses showed strong agreement (>85%) on trusting healthcare professionals’ judgments, receiving adequate information, and perceiving medicines as effective and beneficial. However, substantial burden emerged in practical aspects, with 54.6% disagreeing that medicines do not interfere with daily tasks, 45.4% concerned about side-effect severity, and 20.3% worried about costs. Interference with sexual life (23.6% agreement) and social relationships (16.4% agreement) was also notable (S1 Fig).

**Table 2.**
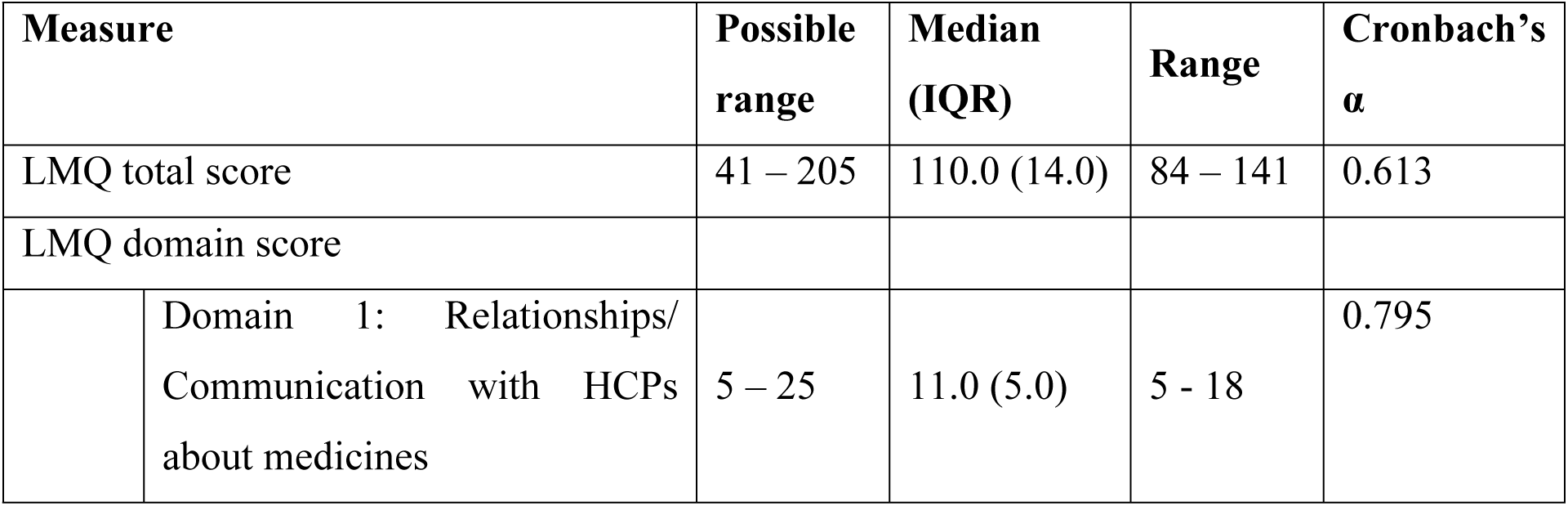
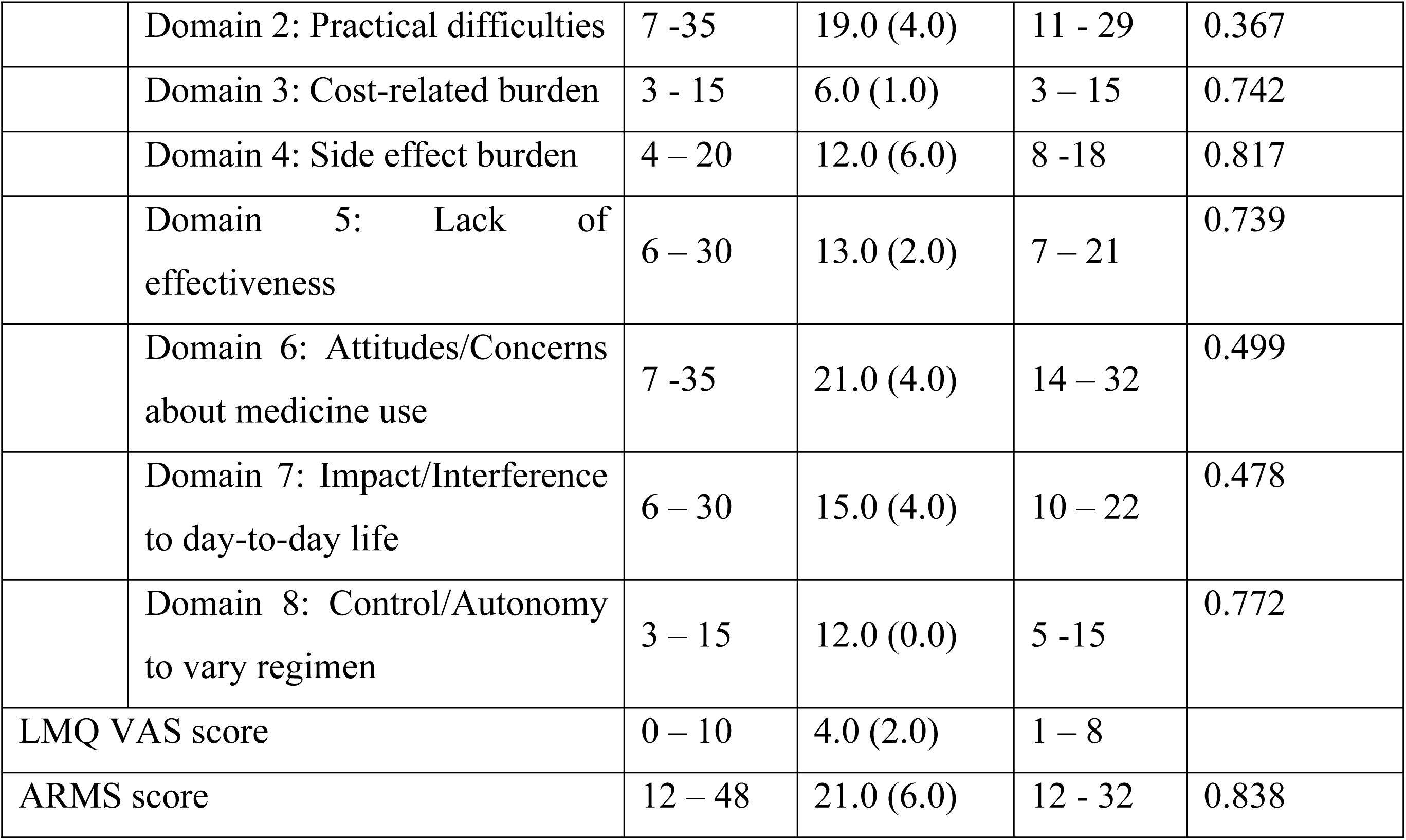
LMQ total score, domain scores and ARMS score of participants (N = 390)

Total LMQ-3 scores were significantly higher among participants with type 2 diabetes mellitus or hypertension as primary diagnosis compared to those with chronic obstructive pulmonary disease or ischemic heart disease (medians 114 vs. 107, p < 0.001), those with multimorbidity compared to without (113 vs. 103, p < 0.001), severe versus mild/moderate Charlson Comorbidity Index categories (116 vs. 106–108.5, p < 0.001), polypharmacy compared to none (114.5 vs. 101.5, p < 0.001), oral-plus-non-oral formulations compared to oral-only (109 vs. 106, p < 0.001), twice-daily compared to once-daily frequency (113 vs. 106, p < 0.001), those requiring assistance compared to independent (117 vs. 108, p < 0.001), and out-of-pocket payers compared to non-payers (112 vs. 109, p < 0.05) (S2 Fig).

### LMQ domain scores

Domain-specific medians reflected varying burden levels: relationships/communication with healthcare professionals about medicines (Domain 1: 11.0, IQR 5.0), practical difficulties (Domain 2: 19.0, IQR 4.0), cost-related burden (Domain 3: 6.0, IQR 1.0), side-effect burden (Domain 4: 12.0, IQR 6.0), lack of effectiveness (Domain 5: 13.0, IQR 2.0), attitudes/concerns about medicine use (Domain 6: 21.0, IQR 4.0), impact/interference to day-to-day life (Domain 7: 15.0, IQR 4.0), and control/autonomy to vary regimen (Domain 8: 12.0, IQR 0.0). Internal consistency (Cronbach’s α) ranged from 0.367 (practical difficulties) to 0.817 (side-effect burden) (Table 2).

Significant domain differences across participant characteristics paralleled total score patterns (S1 Table). For instance, practical difficulties (Domain 2) scores were higher in those with multimorbidity versus those with not (20.0 vs. 18.0, p < 0.05), severe versus mild Charlson Comorbidity Index (21.0 vs. 18.0, p < 0.05), polypharmacy versus none (20.0 vs. 18.0, p < 0.05), twice-daily versus once-daily frequency (20.0 vs. 18.0, p < 0.05), and assistance-requiring versus independent (20.0 vs. 18.0, p < 0.05). Side-effect burden (Domain 4) was elevated in type 2 diabetes mellitus/hypertension versus chronic obstructive pulmonary disease (12.0–13.0 vs. 11.5, p < 0.05), multimorbidity (12.0 vs. 10.0, p < 0.05), severe Charlson Comorbidity Index (13.0 vs. 10.5–11.0, p < 0.05), polypharmacy (12.0 vs. 10.0, p < 0.05), oral-plus-non-oral formulations (12.0 vs. 12.0, but significant per table), twice-daily frequency (12.0 vs. 12.0, but significant), assistance-requiring (14.0 vs. 10.0, p < 0.05), and out-of-pocket payers (12.0 vs. 12.0, but significant). Similar comparative elevations occurred in cost-related burden (Domain 3), impact/interference (Domain 7), and other domains among these groups.

### Predictors of medication-related burden (LMQ Score)

The predictive performance of six machine learning models - Ordinary Least Square Regression (OLS), Penalized Linear Regression (LR), Random Forest (RF), Extreme Gradient Boosting (XGBoost), Light Gradient Boosting Machine (LightGBM), and Support Vector Machines (SVM) – was evaluated (Fig 1A). RF emerged as the most robust model across all primary error metrics, achieving lowest RMSE (7.526, 95% CI [6.277, 8.824]), MAE (5.904, 95% CI [4.846, 7.097]) and Huber loss (5.433, 95% CI [4.387, 6.618]). The RF model also demonstrate the highest variance explained (*R^2^* = 0.540, 95% CI [0.383, 0.685]), outperforming OLS (*R^2^* = 0.476, 95% CI [0.368, 0.513]) and XGBoost (*R^2^* = 0.387, 95% CI [0.205, 0.563]).

**Fig 1.**
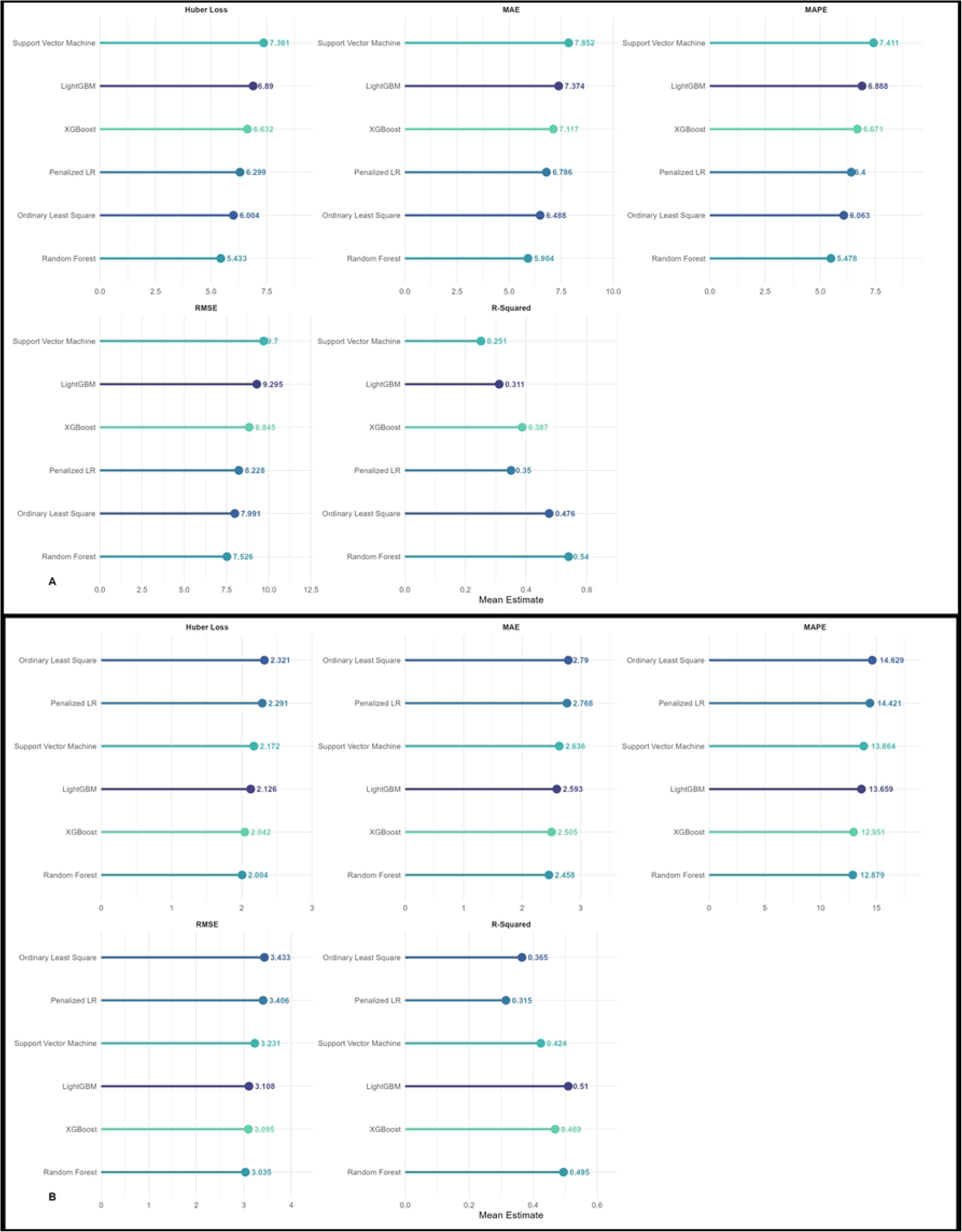
Evaluation of predictive accuracy across six regression architectures. **A. For LMQ Score. B. For ARMS Score** MAE: Mean Absolute Error; MAPE: Mean Absolute Percentage Error; RMSE: Root Mean Square Error

**Fig 2.**
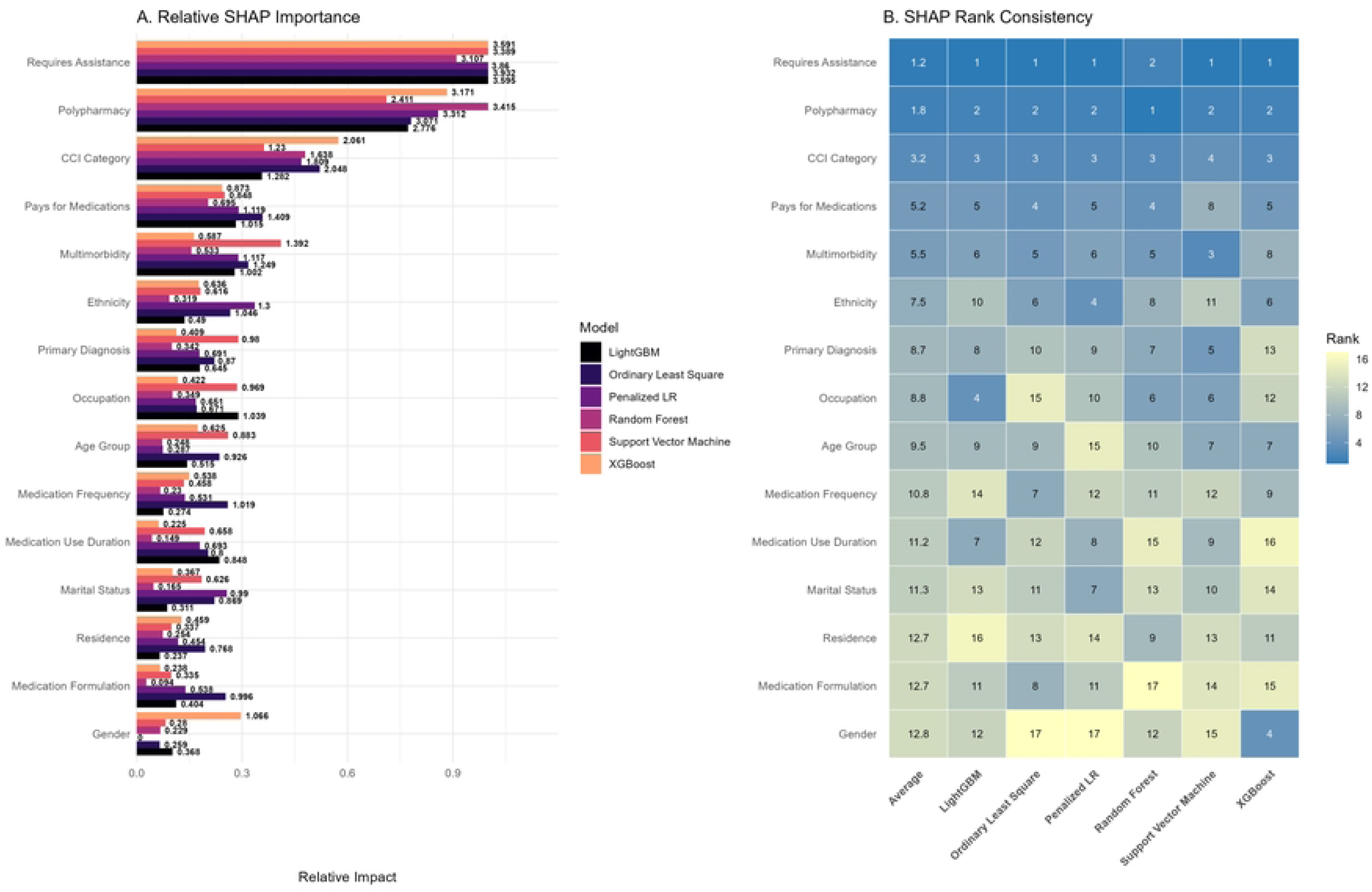
Global interpretability and feature importance consistency across the evaluated models using SHAP (Shapley Additive exPlanations) values to predict LMQ Score. A. Bar-graph comparing SHAP values of the 17 features across six models. B. Heatmap representing the rank of each feature’s importance for each specific model and consensus ranking.

The global SHAP (Shapley Additive exPlanations) analysis revealed a high degree of consensus among the models regarding the primary drivers of medication-related burden. Requires assistance emerged as the most influential predictors, ranking first across five out of six models, closely followed by Polypharmacy. Clinical features (CCI category, Multimorbidity and Primary Diagnosis) were consistent middle-tier predictors. Among medication related variables, Polypharmacy and Pays for Medications were among top-tier predictors. Socio-demographic features (Occupation, Marital Status, Residence, and Gender) showed lowest predictive power across the board.

Multivariate analysis yielded standardized beta (β) values, providing a clear measure of association strength and directionality of each feature (Fig 3A). Consistent with SHAP rankings, Requires assistance and Polypharmacy were the strongest positive predictors of medication-related burden (β = 0.84, β = 0.67, p < 0.05, respectively). The strongest negative predictors were having a moderate CCI (β = −0.39, p < 0.05) and diagnosis with Type 2 Diabetes Mellitus (β = −0.25, p < 0.05).

**Fig 3.**
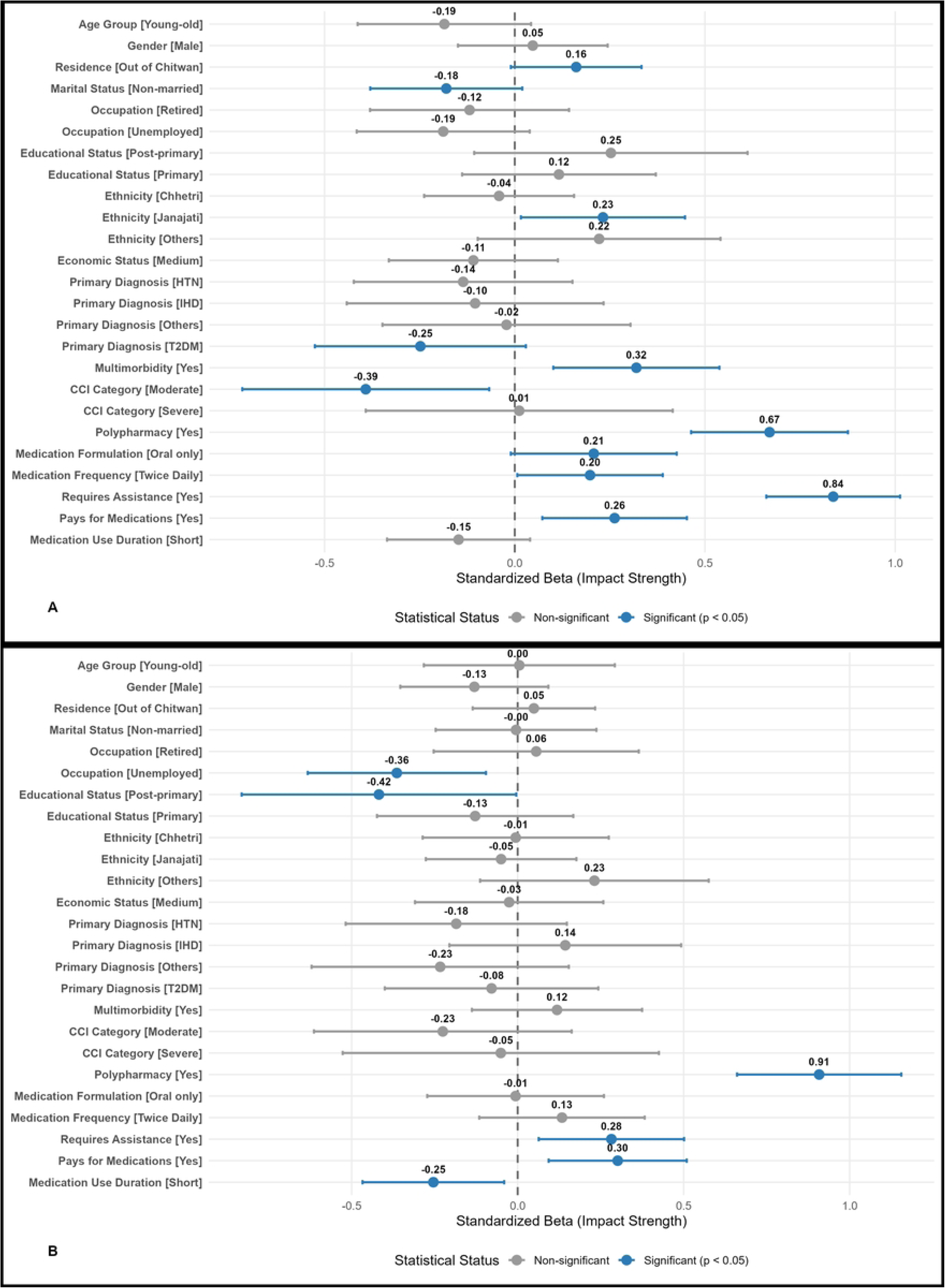
Forest plot of predictors based on OLS regression. A. For LMQ Score. B. For ARMS Score

The SHAP dependence analysis revealed that the RF model’s prediction of the LMQ score is most heavily driven by two dominant factors: Polypharmacy and Requires assistance (Fig 4). Interaction showed that even if one feature is absent, presence of another feature increased the SHAP values, implying increased medication-related burden. The CCI category highlighted a clear interaction effect; while severe comorbidity naturally increased risk, this burden was significantly amplified for patients who also required physical or cognitive assistance. Similarly, medication costs served as a distinct divider, where the financial burden of paying for medications consistently pushed SHAP values higher (implying higher medication-related burden), especially when compounded by requirement of assistance. Multimorbidity showed a consistent step function, moving the risk from a protective negative value to a positive one once present. However, the impact of occupation was more nuanced and of lower magnitude, where being unemployed lowered the medication-related burden. Overall being employed increased the medication-related burden but this effect was somewhat protected by requiring assistance.

**Fig 4.**
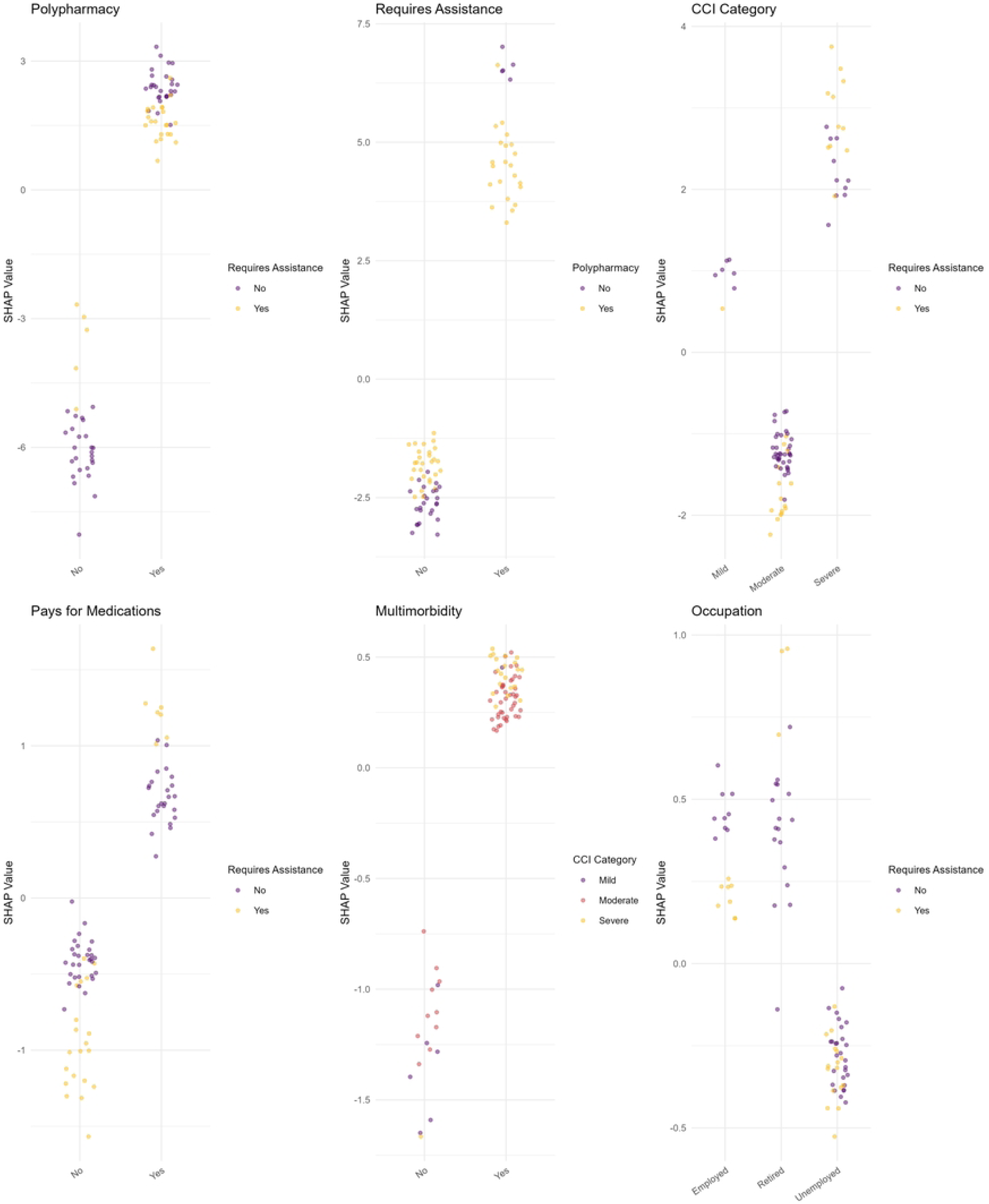
Visualization of marginal effects and feature interaction for the top six predictors of LMQ score in the RF model.

### Medication adherence (ARMS Score)

The median Adherence to Refills and Medications Scale (ARMS) score was 21.0 (IQR 6.0; range 12–32), suggesting moderate non-adherence, with strong reliability (Cronbach’s α=0.838, McDonald’s ω=not reported) (Table 2). Item distributions indicated frequent non-adherence behaviors: 69.5% reported sometimes or never forgetting prescriptions, 63.1% sometimes running out of medicines, and 56.7% sometimes changing doses to suit needs. Better adherence was seen in planning refills ahead (56.9% none of the time) and not missing doses when feeling better (41.5% none) or sick (38.7% none) (S3 Fig).

ARMS scores (higher indicating worse adherence) were significantly elevated in males compared to females (22 vs. 21, p < 0.05), primary-educated compared to illiterate/post-primary (20.5 vs. 21, p < 0.05), type 2 diabetes mellitus or ischemic heart disease versus chronic obstructive pulmonary disease or hypertension (23–24 vs. 19–21, p < 0.001), multimorbidity versus absence of multimorbidity (22 vs. 19, p < 0.001), severe versus mild/moderate Charlson Comorbidity Index (23 vs. 19–21, p < 0.001), polypharmacy versus none (23 vs. 19, p < 0.001), oral-plus-non-oral versus oral-only formulations (23 vs. 21, p < 0.001), twice-daily versus once-daily frequency (23 vs. 19, p < 0.001), assistance-requiring versus independent (22 vs. 21, p < 0.05), and out-of-pocket payers versus non-payers (23 vs. 20, p < 0.001) (S4 Fig).

### Predictors of medication adherence (ARMS Score)

Consistent with findings of LMQ score, RF model demonstrated superior predictive performance for the ARMS score, achieving lowest error rate across all primary metrics, including RMSE (3.035, 95% CI [2.644, 3.472]), MAE (2.458, 95% CI [2.081, 2.879]), and Huber loss (2.004, 95% CI [1.645, 2.411]). While the LightGBM model demonstrated highest variance explained (*R^2^* = 0.510, 95% CI [0.353, 0.649]), RF (*R^2^* = 0.495, 95% [0.342, 0.635]) followed closely, significantly outperforming OLS baseline (*R^2^* = 0.365, 95% CI [0.265, 0.399]). Consistent with LMQ findings, the SVM and LR models generally exhibited higher predictive errors (Fig 1B).

Global interpretability analysis via SHAP values identified Polypharmacy as the most critical and consistent predictor of medication adherence, ranking first across all six evaluated models (Fig 5). Requires assistance, pays for medication, occupation and medication use duration, closely followed as mid-tier predictors. Conversely, demographic variables such as Age group, Residence, Ethnicity and Educational status consistently showed lower predictive impact on medication adherence. Standardized beta coefficients further quantified these clinical effects, revealing that polypharmacy (β = 0.91, p < 0.05) has the strongest positive association with higher ARMS score (indicating poorer adherence), followed by Pays for medications (β = 0.30, p < 0.05) and Requires assistance (β = 0.28, p < 0.05) (Fig 3B). Interestingly, post-primary education status (β = −0.42, p < 0.05) and being unemployed (β = −0.36, p < 0.05) were strongest negative predictors, implying significantly better adherence.

**Fig 5.**
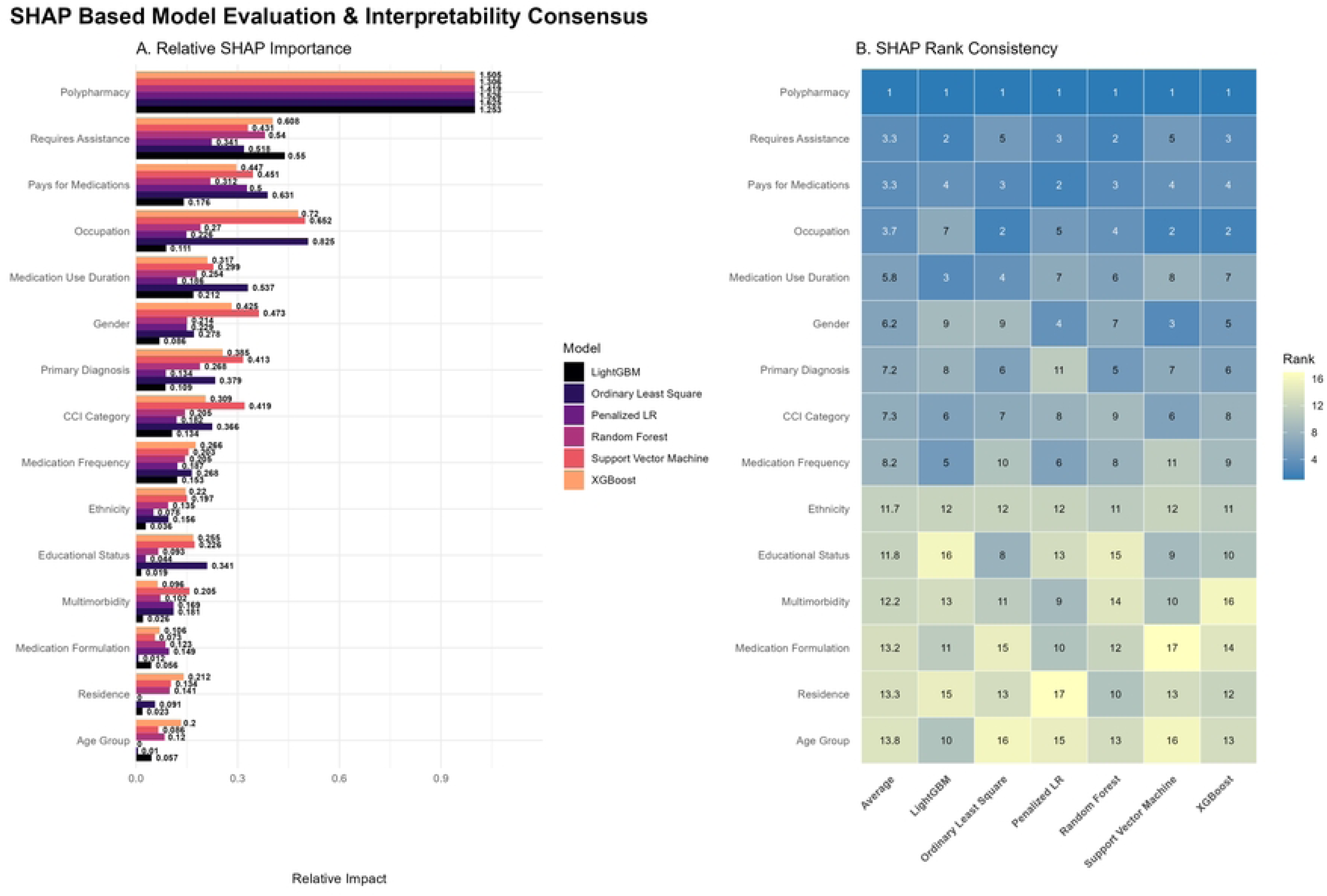
Global interpretability and feature importance consistency across the evaluated models using SHAP (Shapley Additive exPlanations) values to predict ARMS Score. A. Bar-graph comparing SHAP values of the 17 features across six models. B. Heatmap representing the rank of each feature’s importance for each specific model and consensus ranking.

The SHAP dependence analysis revealed that the RF model’s prediction of the ARMS score is also most heavily driven by two dominant factors: Polypharmacy and Requires assistance (Fig 6). A key interaction existed where requiring assistance potentially offsets some of the risk of polypharmacy, whereas needing help with a simpler medication regimen marks a peak risk for poor adherence. Paying for medication and having twice-daily dosing regimen further escalated the risk, as did the long-term duration of medication use, particularly when combined with presence of polypharmacy. Interestingly, being employed increased the risk of medication non-adherence, with polypharmacy further exacerbating these values.

**Fig 6.**
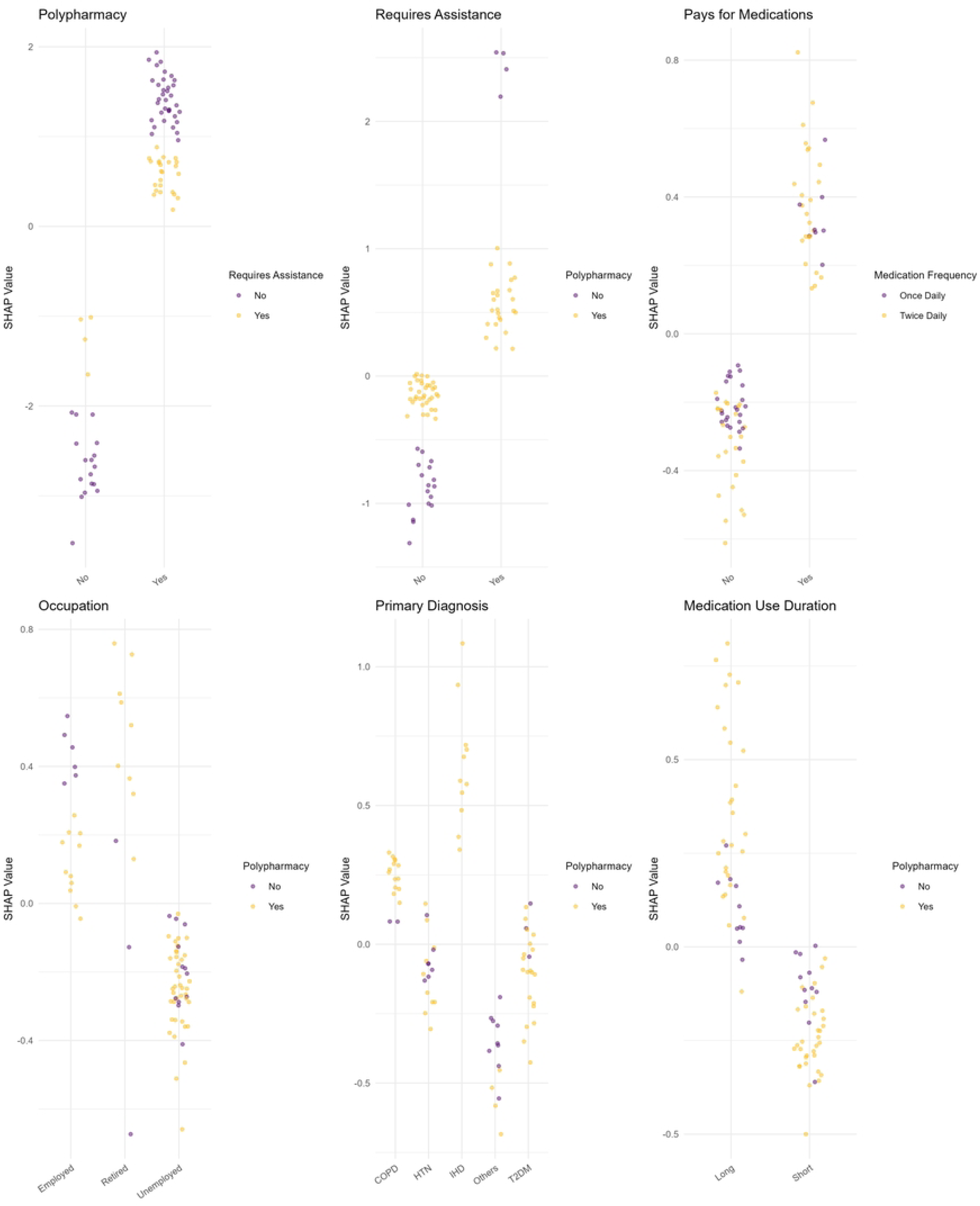
Visualization of marginal effects and feature interaction for the top six predictors of ARMS score in the RF model.

## Discussion

This study represents the first assessment of the medication-related burden from patient perspectives using Living with medicines questionnaire (LMQ-3) in South Asia. The results demonstrate that older adult in Central Nepal experience moderate medication-related burden and moderate non-adherence, By employing machine learning architectures, we identified that requiring assistance for medication management and polypharmacy were the primary drivers for both increased MRB and poor adherence, whereas traditional socio-demographic factors like age and gender were weak predictors.

A key finding of this study was the superior performance of random forest (RF) model in predicting both MRB and adherence, significantly outperforming the OLS regression baseline. This suggests that the relationship between patient characteristics and medication behaviors are non-linear and complex. Traditional statistical methods may overlook these intricate patterns, whereas ML provides a more robust framework for identifying high-risk patients in clinical settings.

Our study found the moderate burden among the participants enrolled in the study which is similar to that of the study done in Kuwait, Qatar but lower than that of the study conducted in England (30–32). Research on medication-related burden using the LMQ-3 remains scarce in South Asia and other low-to-middle-income countries. Most existing data originate from developed or Arabic nations, which may limit the relevance of direct comparisons. Domain specific analysis of MRB showed the highest burden occurred in the “Attitudes/Concerns about medication use”, “Practical difficulties” and “Control/autonomy about medication use” domain. While participants reported the adequate communication and trust in their healthcare professional (Domain 1), they expressed a need for more detailed medication information (Item 9), and desire for greater autonomy in selecting drug brands (Item 8). Furthermore, significant concerns persisted regarding long-term adverse effects (Item 12), and burden of being reliant on medicines (Item 16). These findings suggest that high levels of trust do not necessarily equate to high level of treatment satisfaction, and clinicians should shift to provide individualized care through shared decision-making. Similar burden was reported in the previous study as well (33). Evidence suggests that active patient participation in decision-making and explicit clinician-led conversations about medications improved the patient care (34,35). Furthermore, trust in healthcare system is associated with superior health outcomes, enhanced treatment adherence, and effective management of the health crisis (36). Although participants showed less cost-related burden and more satisfaction with medication effectiveness, practical difficulties leading to MRB were reported. Participants reported challenges in obtaining prescriptions from doctors (Item 1), and getting medicines from the pharmacy (Item 2). These systemic barriers, combined with the physical and cognitive demands of medication routines, highlight the need for simplified regimes and improved pharmacy access for geriatric population.

SHAP analysis identified “requiring assistance” and “polypharmacy” as the most influential predictors of MRB. The findings of aligns with the previous literature (37), which suggests older adults requiring social support for the practical difficulties of using medicine management experience higher levels of burden. Furthermore, rising rates of polypharmacy present significant challenges in geriatrics pharmacotherapy, contributing to prolonged hospital stay and a higher risk of receiving potentially inappropriate medications; ultimately contributing to higher MRB. Interestingly, the results showed that the burden is not uniform across all chronic conditions. Patients diagnosed with Type 2 Diabetes Mellitus reported significantly higher LMQ scores. This may be due to the demanding nature of daily monitoring and the complex dosing schedules often required for metabolic and cardiovascular management (38,39).

The Adherence to Refills and Medications Scale (ARMS) was particularly designed to assess the medication adherence among older people with low literacy (27). This study identified moderate non-adherence among the older population, with a Median score of 21.0 (IQR: 6.0). This level of non-adherence is higher than that reported in similar study from Ethiopia (40). Item-level analysis suggested that this non-adherence was largely driven by behavioral factors rather than physical constraints, similar to finding from a similar study from (41). Participants frequently missed doses due to carelessness, running out of medication or stopping treatment when they feel better. Additionally, forgetfulness was common, particularly regarding multiple-dose regimens and filling prescriptions. The results also indicate a significant lack of proactive planning for medication refills among the participants. Forgetting medication at older age is common because of the physiological changes of the body (42). Evidence suggest targeting the psychological factors along with treatment satisfaction have potential to enhance medication adherence among these vulnerable population (43).

The behavioral patterns seen in medication adherence are reinforced by the SHAP interpretability analysis, which identified polypharmacy as the most critical predictor of medication adherence. The interaction analysis revealed that the complexity of managing multiple medications exacerbates forgetfulness and the likelihood of running out of supplies. Interestingly, while polypharmacy increases the risk of non-adherence, the presence of caregiver can partially mitigate these behavioral lapses. However, the “peak risk” for poor adherence observed in patients with simpler regimens who still required assistance suggests that functional or cognitive decline may override the benefits of a simplified therapy.

The prevalence of polypharmacy and its significant impact a key influencing predictor of both medication-related burden and non-adherence highlight the importance of the patient-centered approaches such as deprescribing for effective polypharmacy management (44). Although, data on the effective implementation of the de-prescribing for adherence as outcome measurement is still insufficient (45), prior research in Nepal indicates that over half of patient were willing to have their medication de-prescribed if possible (5). This suggests a significant opportunity for medication optimization in in resource-limited settings. However, barriers like clinical acceptance of deprescribing, and patient fears regarding symptom relapse persist (46). By establishing the benefits of deprescribing, this study advocates for polypharmacy management as a primary strategy to reduce the overall medication-related burden among older adults.

Although age was a predictor for overall MRB, participants aged 75 years or above reported significantly higher burden regarding practical difficulties. This finding align with research from Bahrain (47), where patients over 75 were more likely to perceive medication as burden. Evidence suggests as patients age, the risk of polypharmacy increases, making pharmacotherapy challenging, a situation often exacerbated by addition of new doses to existing regimens (48). Consequently, physicians and other health professionals must collaborate to implement the strategic intervention aimed at reducing the medication burden MRB in this vulnerable age group (49).

Financial factors also played a significant role. Out-of-pocket payments were positively associated with both higher MRB and poorer adherence. SHAP dependence plots conformed that paying for medication consistently pushed risk values higher, especially when combined with need for assistance. This is particularly relevant in Nepal’s evolving healthcare system, where co-payment structures vary by age and insurance status. Interestingly, higher education (post-primary) and being unemployed were associated with better adherence. The latter may be due to unemployed individuals having more time to manage complex dosing schedules compared to those currently working.

Despite the magnitude of the behavioral problem of non-adherence in older population, existing studies have not found substantial level population based study.

### Strengths and Limitations

The study’s primary strength lies in its use of robust ML architectures, which achieved higher predictive accuracy than traditional linear models by capturing non-linear interactions between variables. The sample size was calculated a prior to ensure statistical power for multivariable modeling. However, there were some limitations. The study was conducted at a single center among patients enrolled in a specific health insurance package, which may limit generalizability of the findings to the broader Nepalese geriatric population. Furthermore, while we followed a rigorous translation process, the internal consistency for certain LMQ-3 domains was low, suggesting that the tool may require further culture validation for the Nepalese context.

## Conclusion

The study established that older adults in Central Nepal experience a moderate level of medication-related burden and non-adherence. The findings identified receiving assistance and polypharmacy as the most significant drivers of both increased medication-relatd burden and poor adherence. The study further revealed that while patients trust healthcare providers, they remain burdened by concerns over long-term side effects and practical management challenges. Behavioral lapses, such as forgetfulness and poor refill planning, further exacerbate non-adherence, particularly among those with complex regimens. Consequently, clinical practice must transition toward patient-centered strategies, including routine medication reviews and deprescribing protocols, to reduce therapeutic complexity and improve health outcomes in resource-limited settings.

## Data Availability

All the relevant data are included in this article. Detailed data, if required, can be available from corresponding author on reasonable requisition.

## Acknowledgement

We would like to acknowledge the original developer of the Living with Medicines Questionnaire (LMQ-3) and Adherence to Medication refills Scale (ARMS) for letting their tool to use in our Study. Further, we would like to acknowledge all the patients, Pharmacy department of the Bharatpur Hospital who contributed to the process data collection for this study.

## Supporting information

**S1 File. Transparent Reporting of a multivariable prediction model for Individual Prognosis or Diagnosis AI (TRIPOD+AI) checklist**

**S2 File. Detailed machine learning pipeline, model optimization and diagnostic framework for predicting medication-related burden and adherence.**

**S3 File. Comprehensive diagnostic evaluation of machine learning models for predicting medication-related burden (LMQ score).**

**S4 File. Comprehensive diagnostic evaluation of machine learning models for predicting medication adherence (ARMS score).**

**S1 Table. LMQ VAS and LMQ domain score distribution across participants characteristics (N = 390)**

**S2 Table. Detailed description of candidate predictor features**

**S1 Fig.**
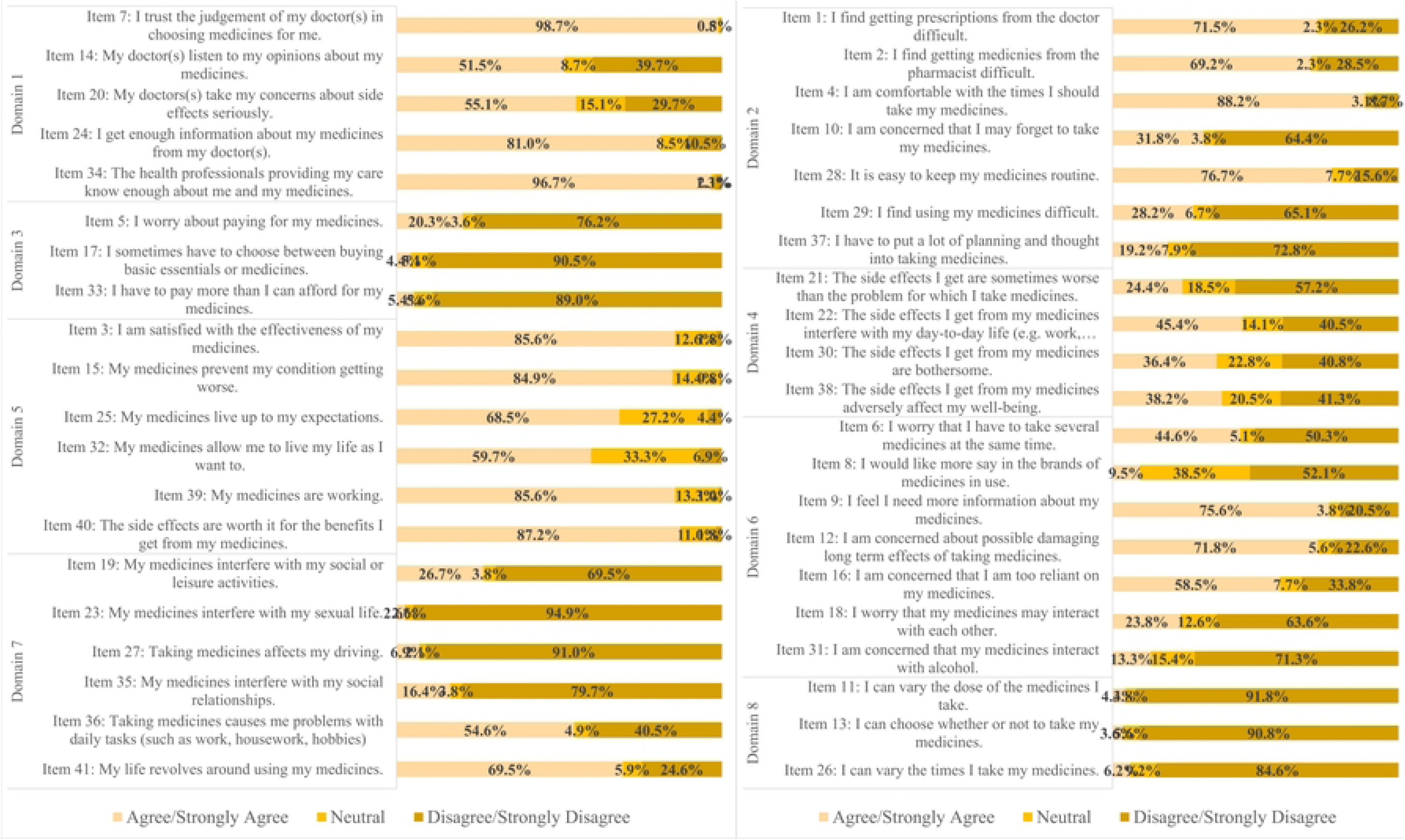
Item-wise distribution of LMQ questionnaire.

**S2 Fig.**
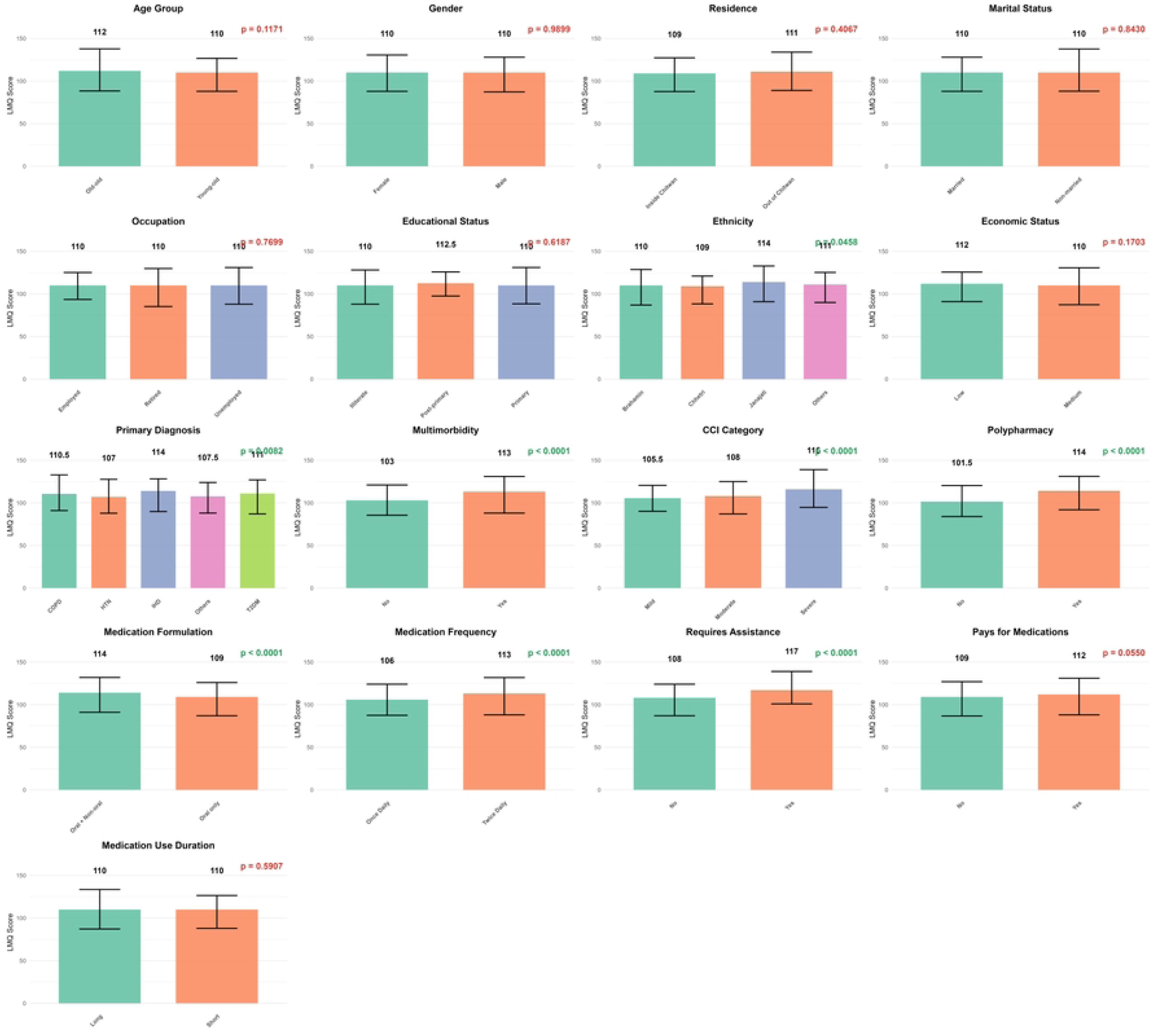
Comparison of Median LMQ scores by participants characteristics.

**S3 Fig.**
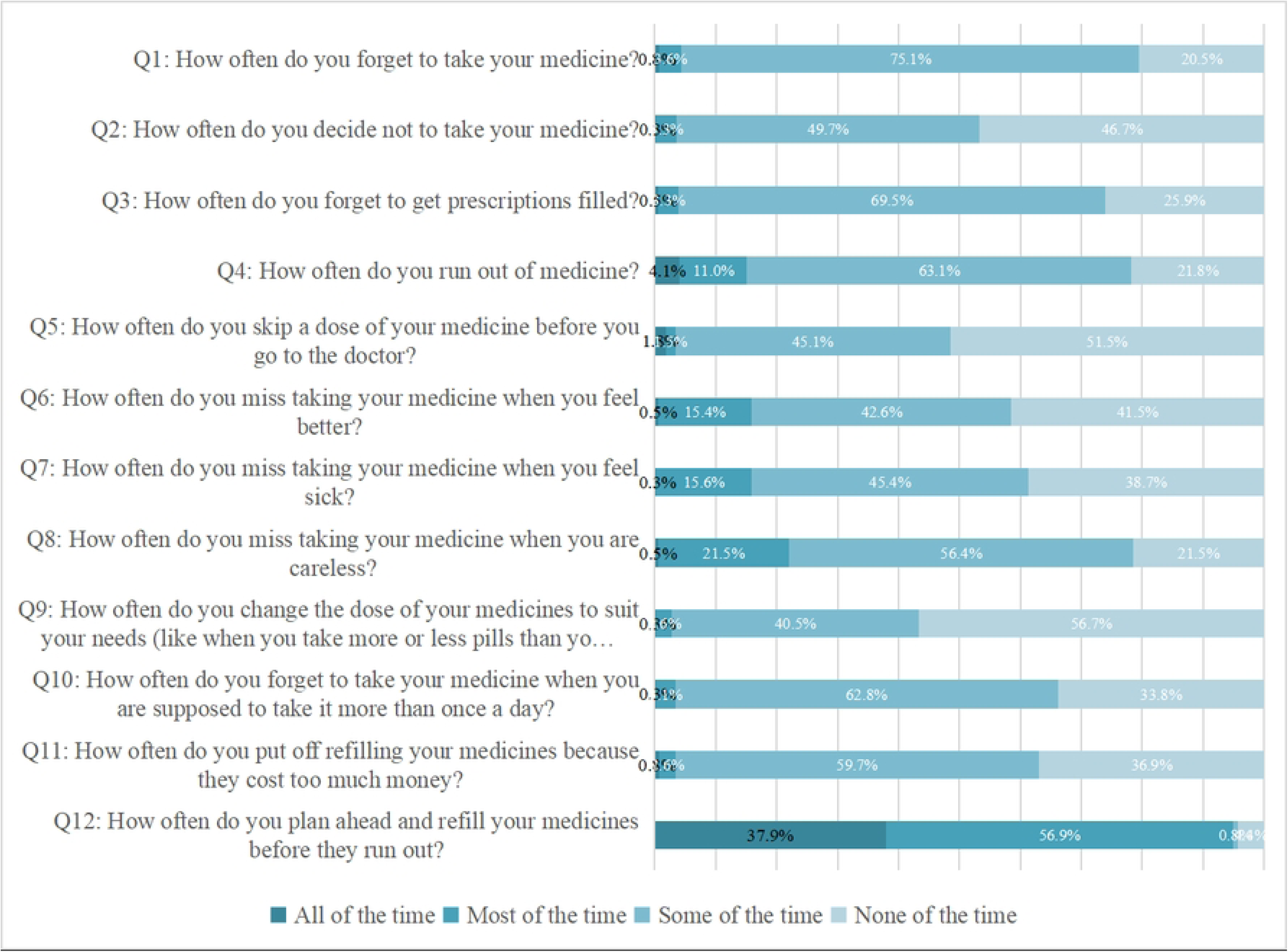
Item-wise distribution of ARMS questionnaire.

**S4 Fig.**
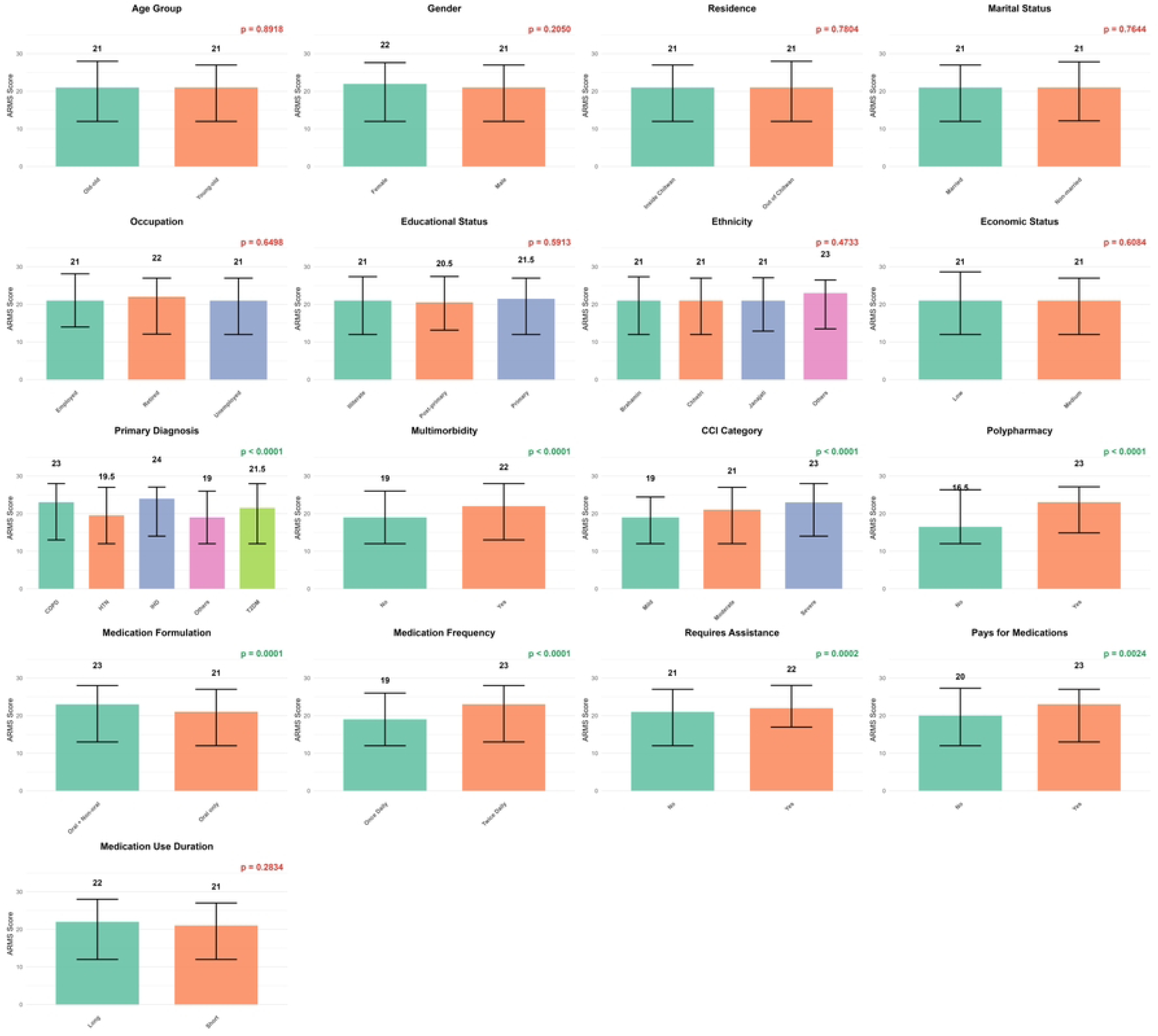
Comparison of Median ARMS scores by participants characteristics.

